# Hypovitaminosis D is associated with sleep disorders in workers on alternating shifts with cardiovascular risk factors

**DOI:** 10.1101/2021.05.04.21256625

**Authors:** Luiz Antônio Alves de Menezes Júnior, Virgínia Capistrano Fajardo, Sílvia Nascimento de Freitas, Fausto Aloísio Pedrosa Pimenta, Fernando Luiz Pereira de Oliveira, George Luiz Lins Machado-Coelho, Raimundo Marques do Nascimento Neto, Adriana Lúcia Meireles

## Abstract

Shift work has serious health impacts due to desynchronization of the circadian rhythm; consequently, the workers have increased sleep disturbances. Another impact is working hours, which can contribute to decreased sun exposure and lead to the development of hypovitaminosis D. Vitamin D has been implicated in extraskeletal functions in many physiological mechanisms, including sleep. Therefore, we aimed to verify the association between sleep parameters and hypovitaminosis D in shift workers with cardiovascular risk. We conducted a cross-sectional study of 82 male rotating shift workers (24-57 years old) with at least one cardiovascular risk factor (such as hyperglycemia, dyslipidemia, abdominal obesity, physical inactivity, hypertension, and smoking). Polysomnography was used to evaluate sleep parameters, while vitamin D levels were measured using a chemiluminescence method. Logistic regression was used to model the association between hypovitaminosis D and sleep parameters after adjustment for relevant covariates. Hypovitaminosis D (< 20 ng/mL) was seen in 30.5% of the workers. Shift workers with hypovitaminosis D had lower sleep efficiency, increased microarousal index, and lower arterial oxygen saturation after adjusting for seasonality, age, and body fat. Therefore, we suggest that hypovitaminosis D is associated with greater sleep disturbances in rotating shift workers with cardiovascular risk factors.

## Introduction

In the United States, sleep disorders have been associated with seven of the fifteen leading causes of death ^1^. In addition to risk factors that have been well described in the literature (such as the male sex, older age, and being overweight) ^2^, there is a strong association between vitamin D levels and an increased risk for sleep disorders ^3^.

Vitamin D refers to a group of fat-soluble molecules that can be ingested via diet and/or produced in the skin by the action of ultraviolet rays in sunlight ^4^. Current evidence indicates that the pleiotropic effects of vitamin D and its metabolites go far beyond bone-mineral metabolism as well as parathyroid gland activity and are connected to other potential areas, primarily linked to sleep ^3,5^.

Hypovitaminosis D, is a global health problem that affects more than one billion people worldwide ^4^. The most described risk groups are the elderly, pregnant women, as well as patients with osteomalacia, rickets, osteoporosis, inflammatory diseases, autoimmune diseases, chronic kidney disease, and obese individuals ^4^. However, few studies have addressed occupational aspects, such as work shift schedules to be a risk factor. There are two basic forms of shift work: i) permanent, in which the worker works all days at the same time, for example, only during the day, or only during the night; ii) alternating or rotating, where workers rotate shifts, that is, everyone must work in the morning, afternoon, or night shifts and they shift between each. They may have a slow or fast rotation ^6^. Rotating shifts is a predominant factor that promotes discontinuation of a worker’s normal feeding and sleep patterns, inducing changes in circadian rhythm, and consequently, hormonal imbalances as well as disruption of normal sleep architecture ^7^. Besides, shift workers are at high risk for hypovitaminosis D, reaching up to 80% prevalence ^8^.

Therefore, it appears that workers on rotating shifts present risk factors for both hypovitaminosis D as well as changes in sleep quality. However, with the current evidence, it is not possible to conclude whether there is a causal and association effect between the two events, or whether the relationship is justified by mediating variables. Furthermore, no studies have evaluated the association between sleep disturbances and vitamin D levels in rotating shift workers. Therefore, in this study, we investigated whether sleep architecture, measured by polysomnography, is associated with hypovitaminosis D in a sample of rotating shift workers from the Iron Quadrangle region, Brazil.

## Methodology

### Design and Participants

A cross-sectional study was performed on male off-road truck operators (aged 24–57 years) who had rotating shifts at an iron ore extraction company located in Iron Quadrangle, Minas Gerais, Brazil, in 2012. The rotating shifts were timed at 6 h, followed by 12 h of rest, lasting from 7 pm to 1 am, 1 am to 7 am, 7 am to 1 pm, and 1 pm to 7 pm. After completing the four-shift cycle, the workers had a day off.

The participants were previously evaluated in a screening study entitled “Metabolic syndrome in mining workers in the state of Minas Gerais, Brazil,” conducted by the Federal University of Ouro Preto to identify the prevalence of cardiovascular risk factors in this population. For this study, the inclusion criteria were male workers with at least one cardiovascular risk factor from the following: hypertension (systolic blood pressure ≥ 130 mmHg or diastolic blood pressure ≥ 85 mmHg) ^9^, hyperglycemia (fasting blood glucose ≥ 100 mg/dL) ^10^, high total cholesterol (≥ 200 mg/dL), high triglycerides (≥ 150 mg/dL), high low-density lipoprotein (≥ 160 mg/dL), low high-density lipoprotein cholesterol (<40 mg/dL) ^11^, abdominal obesity (waist circumference ≥ 90 cm) ^9^, tobacco use, and low level of physical activity (< 600 measured energy total - min/week) ^12^.

Of the 678 rotating shift workers, 524 had a minimum of one cardiovascular risk factor and were invited to undergo a polysomnography (PSG). Of these, 119 showed up for PSG, and 37 workers did not get their vitamin D levels measured. Therefore, at the end of the study, overall, 82 workers underwent a PSG examination and had their vitamin D measured (Figure 1). This study followed reported guidelines dictated by the Strengthening the Reporting of Observational Studies in Epidemiology (STROBE).

**Figure 1.**
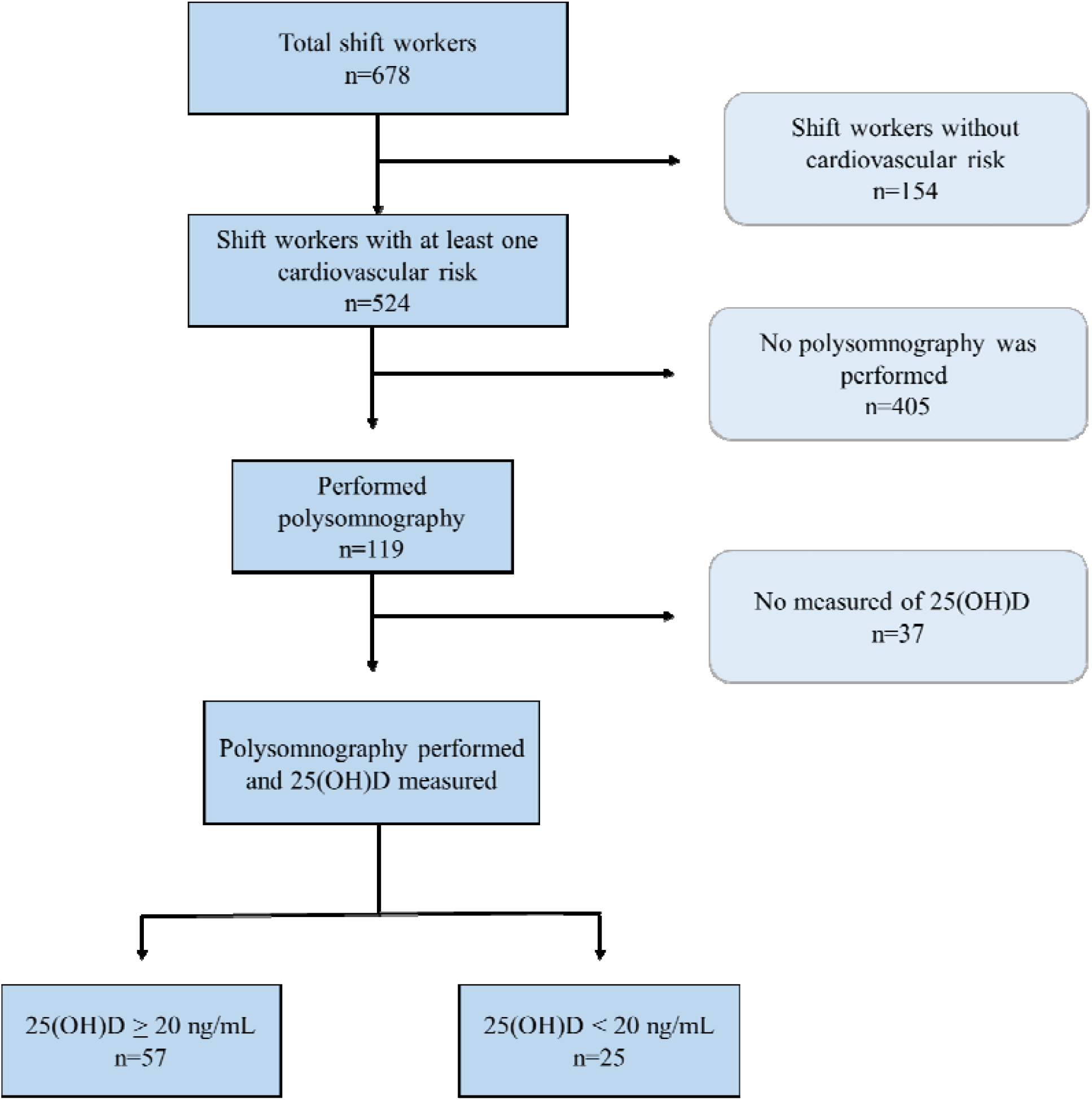
Flowchart of study participants

### Data collection

Data were obtained from the Cardiometabolism Laboratory at the Medical School of the Federal University of Ouro Preto, and a questionnaire administered to the participants. At this time, anthropometric evaluation and blood collection were also performed, while PSG was done on a later day.

The social, demographic, and economic variables evaluated were sex, age, self-reported skin color, and shift work time. Age was categorized as 20-29, 30-39, 40-49, 50-59, and 60 years or more. Overall shift work time was categorized as < 5 or > 5 years. The self-declared skin color was categorized as either being white or not white (black or brown-skinned).

### Polysomnography

All workers underwent a PSG examination at night (10 pm to 6 am) registered using the Alice 5 PSG system (Philips Respironics, Inc., Murrysville, PA, USA) and conducted at Hospital in Ouro Preto.

Sleep parameters evaluated were sleep efficiency, sleep latency, stages of sleep (N1, N2, N3, and REM), REM sleep latency, mean and minimum arterial oxygen saturation (SaO2), a value of SaO2 < 90% during sleep, microarousal index (MAI), and hypopnea apnea index (AHI). Total sleep time was considered as a short duration of fewer than six hours ^13^. Sleep onset latency was classified as increased when greater than 30 min ^13^. Sleep efficiency (%) was calculated as total sleep time divided by total recording time on PSG, and efficiency was classified as reduced if less than 85%.13 The severity of obstructive sleep apnea (OSA) was categorized as no apnea (AHI < 5/h), apnea (AHI > 5/h), moderate to severe (AHI > 15/h), or severe (AHI > 30/h) ^14^. SaO2 was classified as reduced if less than 90% ^14^. Other sleep parameters, such as MAI and sleep stages, were classified according to the 50th percentile (p50).

### Anthropometric data

Weight, body fat, and visceral fat area (VFA) were analyzed using bioelectrical impedance analysis (BIA) on the body composition monitor, InBody 720® (Biospace Co. Ltd. Factory). The percentage of body fat was classified as high if values were above the following in different age groups: 20% in 18 to 35, 27% in 35 to 45, and 28% in 46 to 65 ^15^. VFA was classified with > 130 cm^3^ as excess visceral adipose tissue ^16^. Height was measured using the AlturExata portable stadiometer that had a centimeter-scale with a 1 mm accuracy (AlturaExata, Belo Horizonte, Minas Gerais, Brazil). Body mass index (BMI) was calculated using the following formula: weight (kg) / (height (m))^2^ and classified as overweight if BMI > 25.0 kg/m^2^ or obese if BMI > 30.0 kg/m^2^.

### Biochemical

Fasting glucose, triglycerides, total cholesterol, and high-density lipoprotein (HDL) levels were measured using the enzymatic calorimetric method. Low-density lipoprotein cholesterol (LDL) was calculated using the Friedewald equation (1972) for triglyceride levels less than 400 mg/dL. We classified blood glucose as hyperglycemia when the fasting glucose level was ≥ 100 mg/dL ^17^. Lipid profile was classified as dyslipidemia, when total cholesterol ≥ 190 mg/dL, LDL ≥ 130 mg/dL and/or HDL < 40 mg/dL, and/or triglycerides ≥ 150 mg/dL ^18^.

The concentration of vitamin D [25(OH)D] was measured using a Liaison® chemiluminescence immune analyzer (DiaSorin). The participants were classified as hypovitaminosis D when 25(OH)D levels were less than 20 ng/mL ^19^. The seasonality of blood sample collection was classified as either summer (December 21 to March 19), autumn (March 20 to June 20), or spring (September 21 to December 20).

### Statistical analysis

Statistical analyses were performed using Stata (version 15.0), with a significance level of 5%. The Shapiro-Wilk test was performed to assess data distribution. Continuous data are presented as medians and interquartile ranges (IQR) (p25–p75). Categorical data are presented as numbers and percentages. Data were compared using the Whitney and chi-square tests. A binary logistic regression analysis was performed to investigate whether hypovitaminosis D was associated with sleep parameters. Based on the results of the univariate analysis, variables were entered into multiple logistical models in descending order of statistical significance (stepwise technique). Only variables with p ≤ 0.20, biological plausibility, and epidemiological relevance were considered. The adjusted model included variables with p < 0.05, including total body fat, age, and the seasonality of blood collection.

Sampling power (a posteriori) was performed on proportion and sample size data of similar studies using the G*Power program (version 3.1.9.2). The analysis was performed for the whole sample size, with an alpha level of 0.05 (using a two-tailed test), which produced a statistical power of greater than 0.80.

### Ethical issues

This study was conducted in accordance with the guidelines laid down by the Declaration of Helsinki, and all procedures involving human participants were approved by the Research Ethics Committee of the Federal University of Ouro Preto (CAAE: 39682014.7.0000.5150).

## Results

A total of 82 shift workers underwent PSG and vitamin D level measurements. Of these, 30.5% had hypovitaminosis D (25(OH)D level < 20 ng/mL). Most participants were 20 to 39 years old (70.7%), with the youngest at 24.4 and the oldest at 56.2 years. Most worked for more than five years on a rotating shift (80.5%); of these workers, 69.5% had declared themselves non-white. In those that were evaluated (n=82), 75.6% were overweight, 61.0% had high body fat, and 53.7% had a high VFA (Table 1). Workers with hypovitaminosis D had a 4.5% higher body fat percentage when compared to workers with sufficient levels (28.2% [IQR: 9.0] versus 23.7% [IQR: 5.5]; p = 0.010) (data not shown).

**Table 1.** Characteristics of rotating shift workers according to vitamin D levels.

| Characteristics | All workers<br>(n= 82) | 25(OH)D (ng/mL) |  | *p* * |
| --- | --- | --- | --- | --- |
|  |  | ≥ 20 (n= 57) | < 20 (n= 25) |  |
| Age |  |  |  |  |
| 20 – 39 years, n (%) | 58 (70.7) | 43 (75.4) | 15 (60.0) | 0.157 |
| 40 – 59 years, n (%) | 24 (29.3) | 14 (24.6) | 10 (40.0) |  |
| Shift work |  |  |  |  |
| < 5 years | 16 (19.5) | 11 (19.3) | 5 (20.0) | 0.941 |
| ≥ 5 years | 66 (80.5) | 46 (80.7) | 20 (80.0) |  |
| Race/Skin color |  |  |  |  |
| Not white, n (%) | 57 (69.5) | 39 (68.4) | 18 (72.0) | 0.746 |
| Anthropometric data |  |  |  |  |
| Excess weight, n (%) | 62 (75.6) | 42 (73.7) | 20 (80.0) | 0.540 |
| Obesity, n (%) | 22 (26.8) | 13 (22.8) | 9 (36.0) | 0.215 |
| High BF, n (%) | 50 (61.0) | 33 (57.9) | 17 (68.0) | 0.127 |
| VFA > 130 cm³, n (%) | 44 (53.7) | 28 (49.1) | 16 (64.0) | 0.214 |
| Clinical and behavioral variables |  |  |  |  |
| Hypertension, n (%) | 58 (70.7) | 20 (80.0) | 38 (66.7) | 0.222 |
| <sup>a</sup>Dyslipidemia, n (%) | 45 (54.9) | 28 (49.1) | 17 (68.0) | 0.114 |
| <sup>b</sup>Tobacco consumption, n (%) | 19 (23.2) | 12 (21.0) | 7 (29.2) | 0.484 |
| Low physical activity, n (%) | 21 (25.6) | 13 (22.8) | 8 (32.0) | 0.380 |
| <sup>c</sup>Hiperglycemia, n (%) | 4 (4.9) | 2 (3.51) | 2 (8.0) | 0.385 |
| Seasonality |  |  |  |  |
| Spring, n (%) | 54 (65.8) | 32 (56.1) | 22 (88.0) | 0.003 |
| Summer, n (%) | 27 (33.1) | 25 (43.9) | 3 (12.0) |  |

*p*\* value for Pearson's chi-square test or Fisher's exact test;
<sup>a</sup>Dyslipidemia: total cholesterol ≥ 190 mg/dL, triglycerides 150 ≥ mg/dL, LDL ≥ 130 mg/dL, HDL < 40 mg/dL.
<sup>b</sup>Current smokers or who quit less than six months ago;
<sup>c</sup>Hiperglycemia: fasting glucose > 100 mg/dL. Excess weight: BMI > 25.0 kg/m<sup>2</sup>. Obesity: BMI > 30.0 kg/m<sup>2</sup>.
Abbreviations: VD: Vitamin D; BF: total body fat; VFA: visceral fat area;

Regarding cardiovascular risk factors, 70.7% had hypertension, 54.9% had dyslipidemia, 23.2% had medium to high risk tobacoo intake, 25.6% had a low level of physical activity, and 4.9% had hyperglycemia. Furthermore, vitamin D levels showed seasonal variability, with higher hypovitaminosis D presence during spring (88.0%) than during summer (8.0%) (p < 0.05) (Table 1).

Workers with hypovitaminosis D had a longer sleep latency [(23.0 min; IQR: 20.0) versus (11.5 min; IQR: 11.0); p = 0.016)] and a lower sleep efficiency [(80.9%; IQR: 80.9) versus (86.5%; IQR: 10.0; p= 0.017)] when compared to workers who had higher levels of vitamin D. No other sleep characteristics were significantly different between workers with and without hypovitaminosis D (p > 0.05) (Table 2).

**Table 2.** Sleep parameters of shift workers according to vitamin D levels.

| Characteristics | All workers<br>(n= 82) | 25(OH)D (ng/mL) |  | *p* * |
| --- | --- | --- | --- | --- |
|  |  | ≥ 20 (n= 57) | < 20 (n= 25) |  |
| Sleep features |  |  |  |  |
| Total sleep time (hours) | 6.5 (5.8 – 6.9) | 6.6 (5.9 – 6.9) | 6.2 (5.4 – 6.8) | 0.095 |
| Sleep latency (min) | 13.2 (8.5 – 24.0) | 11.5 (8.5 – 19.5) | 23.0 (8.5 – 28.5) | **0.016** |
| REM sleep latency (min) | 99.7 (79.0 – 147.0) | 100.5 (79.0 – 136.0) | 99.0 (80.0 – 147.0) | 0.705 |
| Sleep efficiency (%) | 85.3 (76.9 – 89.2) | 86.5 (80.0 – 90.0) | 80.9 (73.8 – 86.6) | **0.017** |
| AHI (events/hour) | 13.7 (7.6 – 22.9) | 13.6 (7.9 – 22.3) | 16.3 (6.5 – 26.1) | 0.687 |
| MAI (events/hour) | 8.6 (5.5 – 12.4) | 7.7 (5.3 – 11.7) | 11.3 (7.6 – 13.2) | 0.103 |
| Arterial oxygen saturation |  |  |  |  |
| SaO<sub>2</sub> average (%) | 95.0 (94.0 – 96.0) | 95.0 (94.0 – 96.0) | 96.0 (95.0 – 96.0) | 0.221 |
| SaO<sub>2</sub> minimum (%) | 87.0 (83.0 – 89.0) | 87.0 (84.0 – 89.0) | 86.0 (83.0 – 88.0) | 0.634 |
| Time with SaO<sub>2</sub> < 90%, min | 1.4 (0.1 – 4.8) | 1.4 (0.0 – 7.0) | 1.4 (0.2 – 4.7) | 0.776 |
| Sleep stages |  |  |  |  |
| N1 (% TST) | 5.7 (4.4 – 7.4) | 5.7 (4.3 – 7.2) | 5.3 (4.6 – 7.4) | 0.919 |
| N2 (%TST) | 59.1 (54.0 – 63.7) | 59.7 (53.6 – 64.3) | 56.8 (54.1 – 61.6) | 0.308 |
| N3 (%TST) | 15.8 (11.5 – 19.8) | 14.7 (11.2 – 19.0) | 18.1 (14.7 – 20.3) | 0.323 |
| REM (%TST) | 17.7 (14.5 – 24.6) | 18.5 (13.8 – 24.6) | 17.3 (15.8 – 23.0) | 0.875 |

Results are presented as median (interquartile range);
p\* value for Mann Whitney test.
Abbreviations: VD: vitamin D; MAI: microarousal index; AHI, apnea-hypopnea index; OSA: obstructive sleep apnea; REM: rapid eye movement; TST: total sleep time.

Table 3 shows the association of hypovitaminosis D with the outcomes of sleep parameters evaluated, in univariate and multivariate analysis adjusted for seasonality, age, and total body fat. Among the sleep parameters evaluated, only sleep efficiency and MAI showed significant association with hypovitaminosis D in univariate analysis. In the adjusted model, workers with hypovitaminosis D had 3.68 times the chance of having reduced sleep efficiency (p = 0.016), 5.35 times the chance of having SaO2 < 90% (p = 0.020), and 3.85 times the chance of having increased MAI (p = 0.018) (Table 3).

**Table 3.** Multiple logistic regression analysis of sleep characteristics associated with hypovitaminosis D in shift workers.

| Outcomes | Category | n (%) | 25(OH)D < 20 ng/mL |  |  |  |
| --- | --- | --- | --- | --- | --- | --- |
|  |  |  | OR<sub>crude</sub> | *p* | OR<sub>adjusted</sub> | *p* * |
| Total sleep time | > 6 hours | 56 (68.3) | 1.00 |  | 1.00 |  |
|  | ≤ 6 hours | 26 (31.7) | 1.71 | 0.288 | 1.63 | 0.358 |
| Sleep latency | < 30 min | 70 (85.4) | 1.00 |  | 1.00 |  |
|  | ≥ 30 min | 23 (14.6) | 2.68 | 0.121 | 2.94 | 0.098 |
| Sleep efficiency | > 85% | 56 (68.3) | 1.00 |  | 1.00 |  |
|  | ≤ 85% | 26 (31.7) | 4.41 | 0.005 | 3.68 | **0.016** |
| Sleep apnea | AHI < 5/h | 11 (13.4) | 1.00 |  | 1.00 |  |
|  | AHI 5-15/h | 34 (41.5) | 0.30 | 0.073 | 0.25 | 0.175 |
|  | AHI 15-30/h | 27 (32.9) | 1.48 | 0.407 | 1.56 | 0.295 |
|  | AHI ≥ 30/h | 10 (12.2) | 0.97 | 0.971 | 0.98 | 0.239 |
| Arterial oxygen saturation | ≥ 90% | 20 (24.4) | 1.00 |  | 1.00 |  |
|  | < 90% | 62 (75.6) | 4.18 | 0.041 | 5.35 | **0.020** |
| Microarousal index | < P50 | 42 (51.2) | 1.00 |  | 1.00 |  |
|  | ≥ P50 | 40 (48.8) | 2.72 | 0.048 | 3.85 | **0.018** |
| Sleep stage N1 | ≥ P50 | 42 (51.2) | 1.00 |  | 1.00 |  |
|  | < P50 | 40 (48.8) | 1.20 | 0.699 | 1.20 | 0.290 |
| Sleep stage N2 | ≥ P50 | 42 (51.2) | 1.00 |  | 1.00 |  |
|  | < P50 | 40 (48.8) | 1.92 | 0.177 | 1.92 | 0.180 |
| Sleep stage N3 | ≥ P50 | 42 (51.2) | 1.00 |  | 1.00 |  |
|  | < P50 | 40 (48.8) | 0.60 | 0.294 | 0.69 | 0.571 |
| Sleep stage REM | ≥ P50 | 41 (50.0) | 1.00 |  | 1.00 |  |
|  | < P50 | 41 (50.0) | 1.78 | 0.233 | 1.86 | 0.156 |

*p*\*: value for the Wald test.
OR<sub>crude</sub>: value estimated by the univariate analysis; OR<sub>adjusted</sub>: value adjusted for age, body fat, and seasonality.
Abbreviations: OR: odds ratio; P50: 50th percentile

## Discussion

In this study of rotating shift workers with cardiovascular risk factors, we found a high prevalence of hypovitaminosis D with a significant association of lower sleep efficiency, increased MAI, and SaO2 during sleep.

The prevalence of hypovitaminosis D found in the study was 30.5%, lower than that found in a systematic review by Sowah et al. (2017), where 80% of the shift workers had 25(OH)D value less than 20 ng/mL ^8^. However, in the same review, out of all 71 articles evaluated, only two were from Brazil. The two articles did not evaluate drivers or truck operators, and workers on rotating shifts, which made it difficult to compare the data. Previous studies have suggested that a higher prevalence of hypovitaminosis D is due to the lack of sunlight exposure in shift workers ^20^. But, in contrast, a rotating work schedule allows the worker to also have day shift periods (e.g., from 7 am to 1 pm and from 1 am to 7 am), or time off, which may increase their sun exposure at these times.

Another important issue to highlight is the high incidence of sunlight in Brazil during almost all seasons of the year, contributing to a higher cutaneous synthesis of vitamin D ^21^. However, it is important to note that these workers were evaluated during the spring and summer season, which had a higher incidence of sunlight, which in turn may suggest a higher prevalence of hypovitaminosis D during fall and winter. Although Brazil has a high incidence of sunlight, since its a tropical country, the Iron Quadrangle region is a cold region with altitudes between 1200 to 2000m. Attributed to the cold weather, the workers may increase the amount of clothing that covers more parts of the body to keep warm, thus reducing the exposure of the sunlight on the epidermis during autumn and winter. Our results were similar to those found by Eloi et al. (2016), in which 33.3% of the Brazilian population had vitamin D values less than 20 ng/mL ^22^. Moreover, unlike previous studies that found lower vitamin D levels in younger participants ^23^, our study did not find any significant difference concerning age. The younger age range of the study participants may have interfered, with 96.3% of the sample aged 20 to 49 years. Aging leads to a decrease in concentrations of the vitamin D precursor (7-dehydrocholesterol) in the skin, which in turn decreases its endogenous synthesis from sunlight ^23^. Other variables that were not associated with significant differences in serum vitamin D levels were skin color, tobacco use, and chronic comorbidities (diabetes, hypertension, and obesity).

Of the workers evaluated, 61.0% had high total body fat, and those with hypovitaminosis D had 4.5% more body fat than workers with sufficient levels of the vitamin, suggesting a relationship between total body fat and vitamin D levels. The association of hypovitaminosis D in individuals with excess adiposity has been well described in the literature ^24^. Among the hypotheses that addressed this relationship, the most important one is the trapping of vitamin D in the adipose tissue, due to it being highly fat-soluble ^25^. Besides, inadequate dietary habits are also common in obese individuals, leading to decreased vitamin D intake as well as low sun exposure as a result of physical inactivity ^24^. Finally, inflammatory cytokines, elevated in cases of obesity, are known to inversely affect the bioavailability of vitamin D and also increase its clearance ^26^.

In this study, sleep assessment was conducted overnight using PSG. Rotating shift workers with hypovitaminosis D were 3.68 times more likely to have a sleep efficiency lower than 85% when compared to those with higher levels of the vitamin. A similar result was found by Massa et al. (2015) in 3,048 men where lower vitamin D levels (20 – 30 ng/mL) were associated with a higher likelihood of low sleep efficiency (< 70%) as measured by actigraphy (OR, 1.45; p = 0.004) ^27^. However, the results in the literature remain controversial as Lee et al. (2020), who evaluated the sleep of night and day workers by actigraphy, did not find any association between hypovitaminosis D and reduced sleep efficiency ^28^. However, vitamin D levels < 10 ng/mL were used as a cutoff point, which was different from our study (vitamin D levels < 20 ng/mL). Moreover, workers who were on rotating shifts were not evaluated, a factor that is associated more with poor sleep quality when compared to other shifts ^28^.

Another sleep disorder that is prevalent in the population is the OSA ^2^. Of the rotating shift workers, 86.6% had OSA to some degree but had no association with hypovitaminosis D. The prevalence of OSA found in this study was much higher than that found in a systematic review article by Sakamoto et al. (2018), in which the prevalence in shift workers ranged from 14.3 to 38.1% ^29^. However, it is important to note that our sample was composed entirely of male workers on rotating shifts with at least one cardiovascular risk factor, which is associated with a higher prevalence of OSA. Similar values were found in other studies with individuals at high risk for OSA, 83% in patients with suspected apnea ^30^, and 76% in patients at risk for coronary heart disease ^31^.

Although we found no association between hypovitaminosis D and OSA, we did find that workers with vitamin D levels less than 20 ng/mL were more likely to have a higher MAI (OR: 3.85; p = 0.018) and SaO2 < 90% during sleep (OR: 5.34; p = 0.020). MAI and SaO2 during sleep are known to be directly related to OSA. During sleep, the ventilatory control system is subjected to instabilities, increasing the occurrence of apneas or hypopneas, both of which occur when there is a complete or partial respiratory arrest. This results in a reduction of respiratory flow and a decrease in oxyhemoglobin saturation, either accompanied or not by microarousals ^2^. These results are supported in the study by Goswami et al. (2016), wherein after adjusting for risk factors (age, body mass index, neck circumference, and hypertension), there were no associations between vitamin D levels and OSA (OR: 1.05; 95% CI: 0.72 - 1.52)] ^32^. However, these results remain divergent in the literature. A systematic review of 14 studies with 4937 individuals by Neighbors et al. (2018) demonstrated that serum vitamin D levels were lower in individuals with OSA than those without (mean differences were 2.7% for mild OSA, 10.1% for moderate, and 17.4% for severe ^33^. Alternatively, Archontogeorgis et al. (2018) found vitamin D levels to be lower in patients with OSA and negatively correlated with oxygen desaturation index (r = -0.234, p = 0.011) and percentage of time with oxyhemoglobin saturation < 90% (r = -0.172, p = 0.041) ^34^. These results were also found in a cohort study, where chronic pulmonary disease patients had a higher risk of having hypovitaminosis D (OR, 2.32; 95% CI, 1.43 – 3.75), while the relationship between lung function and systemic vitamin D levels was almost linear even after adjustment for numerous confounders ^35^.

We found hypovitaminosis D to be related to several sleep parameters of rotating shift workers, which remained significant even after adjusting for confounding variables. This association may be explained by the intracellular distribution of vitamin D receptors in areas of the brain that regulate the sleep-wake cycle or through pro-inflammatory mediators. Experimental studies have shown that sleep regulatory substances, such as the tumor necrosis factor-alpha (TNF-α), interleukin-6 (IL-6), and interleukin-1 (IL-1) are inversely related to vitamin D level ^36^. Furthermore, vitamin D can negatively regulate cyclooxygenase-2, an enzyme that controls the production rate of prostaglandin D2 (PGD2) (an important regulator of the sleep-wake cycle) implying that hypovitaminosis D may increase circulating PGD2, contributing to the occurrence of sleep Disorders ^37^. Vitamin D is also involved in the production of melatonin, an essential hormone in the regulation of circadian rhythms and sleep. Melatonin synthesis is controlled by the active form of vitamin D (1,25(OH)D), from the enzyme tryptophan hydroxylase ^38^. This suggests a likely role for vitamin D deficiency in sleep disturbances ^36^. These results are corroborated by intervention studies but remain controversial ^39,40^.

Therefore, we can suggest that hypovitaminosis D can negatively affect the quality of sleep in rotating staff workers, leading to an increase in daytime sleepiness, which may impair daily activities. Among these consequences, impaired performance of professional activities, impaired social relationships, and altered cognitive performance can result in an increased risk of work and/or road accidents. This is especially true for the population of workers who are machine operators.

This study is the first to evaluate shift workers from a mining company, specifically truck drivers, who may be at risk for sleep problems. We have shown that hypovitaminosis D is associated with sleep disorders as assessed by polysomnography, the gold standard method of sleep assessment. However, a limitation for the study could be the selection of participants who had at least one cardiovascular risk factor, which limits extrapolation to different populations. Moreover, working in shifts results in an altered circadian cycle rhythm increases metabolic alterations with a higher risk of developing hyperglycemia, dyslipidemia, and hypertension. Besides, altered work schedules can also lead shift workers to have increased physical inactivity.

## Conclusion

Rotating shift workers with cardiovascular risk factors have a high prevalence of hypovitaminosis D and sleep disturbances. We found pertinent results regarding the association of hypovitaminosis D with a longer microarousal index, lower sleep efficiency, and lower SaO2 during sleep. This could lead to increased daytime sleepiness and may interfere with the daily life as well as the work activities of these workers. Further research is expected to elucidate whether changes in vitamin D levels may influence sleep quality in shift workers, in conjunction with clinical trials of using vitamin D supplementation in reducing these parameters.

## Data Availability

The data that support the findings of this study are available on request from the corresponding author, LAAM. The data are not publicly available due to restrictions their containing information that could compromise the privacy of research participants.

## Contributors

VCF, RMNN, SNF, FLPO, FAPP and GLLM contributed to the conception and design of the work, to the acquisition, analysis, and interpretation of data, and the draft of the manuscript. LAAMJ and ALM contributed to the analysis of the results and to the writing of the manuscript. All authors have approved the submitted version.

## Funding

This study was funded by CNPq-Brazil, professor’s productivity financial support and CAPES-Brazil, for PhD student scholarship.

## Competing interests

This was not an industry-supported study. The authors have indicated no financial conflicts of interest. There is no off-label or investigational use in this study.

## Notes

### Competing Interest Statement

The authors have declared no competing interest.

### Funding Statement

This study was funded by CNPq-Brazil, professors productivity financial support and CAPES-Brazil, for PhD student scholarship.

## References

1. Kochanek KD, Murphy S, Xu J, Arias E. Mortality in the United States, 2016. NCHS Data Brief [Internet]. 2017;(293):1–8. Available from: http://www.ncbi.nlm.nih.gov/pubmed/29319473

2. Lee W, Nagubadi S, Kryger MH, Mokhlesi B. Epidemiology of obstructive sleep apnea: A population-based perspective. Vol. 2, Expert Review of Respiratory Medicine. Taylor & Francis; 2008. p. 349–64.

3. Gao Q, Kou T, Zhuang B, Ren Y, Dong X, Wang Q. The Association between Vitamin D Deficiency and Sleep Disorders: A Systematic Review and Meta-Analysis. Nutrients [Internet]. 2018 Oct 1 [cited 2019 Nov 5];10(10):1395. Available from: /pmc/articles/PMC6213953/?report=abstract

4. Hossein-nezhad A, Holick MF. Vitamin D for Health: A Global Perspective. Mayo Clin Proc [Internet]. 2013 Jul 1 [cited 2018 Jun 19];88(7):720–55. Available from: http://linkinghub.elsevier.com/retrieve/pii/S0025619613004047

5. Lai Y-H, Fang T-C. The Pleiotropic Effect of Vitamin D. ISRN Nephrol [Internet]. 2013 Sep 4;2013:1–6. Available from: https://www.hindawi.com/journals/isrn/2013/898125/

6. Simões Mrl, Marques FC, Rocha A de M. Work in Rotating Shifts and its Effects on the Daily Life of Grain Processing Workers. Rev Lat Am Enfermagem [Internet]. 2010 Dec [cited 2018 Jun 19];18(6):1070–5. Available from: http://www.scielo.br/scielo.php?script=sci_arttext&pid=S0104-11692010000600005&lng=en&tlng=en

7. Johnston JD, Ordovás JM, Scheer FA, Turek FW. Circadian Rhythms, Metabolism, and Chrononutrition in Rodents and Humans. Adv Nutr. 2016 Mar 1;7(2):399–406.

8. Sowah D, Fan X, Dennett L, Hagtvedt R, Straube S. Vitamin D levels and deficiency with different occupations: a systematic review. BMC Public Health [Internet]. 2017 Dec 22 [cited 2018 Jun 19];17(1):519. Available from: http://www.ncbi.nlm.nih.gov/pubmed/28637448

9. Alberti Kgmm, Zimmet P, Shaw J. Metabolic syndrome - A new world-wide definition. A consensus statement from the International Diabetes Federation. Vol. 23, Diabetic Medicine. 2006. p. 469–80.

10. American Diabetes Association. Diagnosis and Classification of Diabetes Mellitus. Diabetes Care [Internet]. 2008 Jan 1 [cited 2020 Apr 13];31(Supplement 1):S55–60. Available from: http://www.ncbi.nlm.nih.gov/pubmed/18165338

11. Xavier HT, Izar MC, Faria Neto JR, Assad MH, Rocha VZ, Sposito AC, et al. V Diretriz Brasileira de Dislipidemias e Prevenção da Aterosclerose. Arq Bras Cardiol [Internet]. 2013 [cited 2019 Dec 7];101(4):01–22. Available from: www.arquivosonline.com.br

12. IPAQ RC. Guideline for Data Processing and Analysis of the International Physical Activity Questionnaire (IPAQ): short and long forms. 2005;1–15.

13. Schutte-Rodin SL, Broch L, Buysee D, Dorsey C, Sateia M. Clinical guideline for the evaluation and management of chronic insomnia in adults. J Clin Sleep Med. 2008;4(5):487–504.

14. Thorpy M. International classification of sleep disorders. In: Sleep Disorders Medicine: Basic Science, Technical Considerations and Clinical Aspects: Fourth Edition. Springer New York; 2017. p. 475–84.

15. Pollock ML, Wilmore JH. Exercício na Saúde e na Doença: Avaliação e Prescrição para Prevenção e Reabilitação. 2nd ed. Medsi, editor. São Paulo; 1993. 1–487 p.

16. Després J-P, Lamarche B. Effects of Diet and Physical Activity on Adiposity and Body Fat Distribution: Implications for the Prevention of Cardiovascular Disease. Nutr Res Rev [Internet]. 1993 Jan [cited 2020 Jul 10];6(1):137–59. Available from: https://pubmed.ncbi.nlm.nih.gov/19094306/

17. Oliveira JEP, Montenegro Junior RM, Vencio S. Diretrizes Sociedade Brasileira de Diabetes 2017-2018. 2017.

18. Précoma DB, de Oliveira Gmm, Simão AF, Dutra OP, Coelho OR, Izar MC de O, et al. Updated cardiovascular prevention guideline of the Brazilian society of cardiology – 2019. Arq Bras Cardiol. 2019;113(4):787–891.

19. de Moraes Acf, Maeda SS, Batista MC, Lazaretti-Castro M, Vasconcellos L de S, Madeira M, et al. Posicionamento Oficial da Sociedade Brasileira de Patologia Clínica/ Medicina Laboratorial e da Sociedade Brasileira de Endocrinologia e Metabologia. J Bras Patol Med Lab [Internet]. 2018;53(6):377–81. Available from: http://bibliotecasbpc.org.br/arcs/pdf/PosicionamentoOficial_SBPCML_SBEM_2018.pdf

20. Coppeta L, Papa F, Magrini A. Are shiftwork and indoor work related to D3 Vitamin deficiency? A systematic review of current evidences. Vol. 2018, Journal of Environmental and Public Health. 2018.

21. Mendes MM, Hart KH, Botelho PB, Lanham-New SA. Vitamin D status in the tropics: Is sunlight exposure the main determinant? Nutr Bull [Internet]. 2018 Dec 1 [cited 2020 Jul 18];43(4):428–34. Available from: http://doi.wiley.com/10.1111/nbu.12349

22. Eloi M, Horvath D V., Szejnfeld VL, Ortega JC, Rocha DAC, Szejnfeld J, et al. Vitamin D deficiency and seasonal variation over the years in São Paulo, Brazil. Osteoporos Int [Internet]. 2016;27(12):3449–56. Available from: http://dx.doi.org/10.1007/s00198-016-3670-z

23. Meehan M, Penckofer S. The Role of Vitamin D in the Aging Adult. J Aging Gerontol [Internet]. 2014 Dec 31 [cited 2020 Jul 18];2(2):60–71. Available from: /pmc/articles/PMC4399494/?report=abstract

24. Vanlint S, Simon. Vitamin D and Obesity. Nutrients [Internet]. 2013 Mar 20 [cited 2018 Jun 18];5(3):949–56. Available from: http://www.mdpi.com/2072-6643/5/3/949

25. Savastano S, Barrea L, Savanelli MC, Nappi F, Di Somma C, Orio F, et al. Low vitamin D status and obesity: Role of nutritionist. Rev Endocr Metab Disord. 2017;

26. Wortsman J, Matsuoka LY, Chen TC, Lu Z, Holick MF. Decreased bioavailability of vitamin D in obesity. Am J Clin Nutr [Internet]. 2000 Sep 1;72(3):690–3. Available from: https://academic.oup.com/ajcn/article/72/3/690/4729361

27. Massa J, Stone KL, Wei EK, Harrison SL, Barrett-Connor E, Lane NE, et al. Vitamin D and Actigraphic Sleep Outcomes in Older Community-Dwelling Men: The MrOS Sleep Study. Sleep [Internet]. 2015 Feb 1 [cited 2020 Jul 17];38(2):251–7. Available from: https://academic.oup.com/sleep/article/38/2/251/2416956

28. Niu S-F, Miao N-F, Liao Y-M, Chi M-J, Chung M-H, Chou K-R. Sleep Quality Associated With Different Work Schedules: A Longitudinal Study of Nursing Staff. Biol Res Nurs [Internet]. 2017 Jul 15;19(4):375–81. Available from: http://journals.sagepub.com/doi/10.1177/1099800417695483

29. Sakamoto YS, Porto-Sousa F, Salles C. Prevalência da apneia obstrutiva do sono em trabalhadores de turno: uma revisão sistemática. Cien Saude Colet [Internet]. 2018 Oct 1 [cited 2020 May 4];23(10):3381–92. Available from: http://www.scielo.br/scielo.php?script=sci_arttext&pid=S1413-81232018001003381&lng=pt&tlng=pt

30. Vaz AP, Drummond M, Caetano Mota P, Severo M, Almeida J, Carlos Winck J. Tradução do Questionário de Berlim para língua Portuguesa e sua aplicação na identificação da SAOS numa consulta de patologia respiratória do sono. Rev Port Pneumol [Internet]. 2011 Mar [cited 2019 Dec 7];17(2):59–65. Available from: http://dx.doi.org/10.1016/S0873-2159(11)70015-0

31. Sorajja D, Gami AS, Somers VK, Behrenbeck TR, Garcia-Touchard A, Lopez-Jimenez F. Independent association between obstructive sleep apnea and subclinical coronary artery disease. Chest. 2008 Apr;133(4):927–33.

32. Goswami U, Ensrud KE, Paudel ML, Redline S, Schernhammer ES, Shikany JM, et al. Vitamin D concentrations and obstructive sleep apnea in a multicenter cohort of older males. Ann Am Thorac Soc [Internet]. 2016 May 1 [cited 2020 Jun 10];13(5):712–8. Available from: http://www.atsjournals.org/doi/10.1513/AnnalsATS.201507-440OC

33. Neighbors CLP, Noller MW, Song SA, Zaghi S, Neighbors J, Feldman D, et al. Vitamin D and obstructive sleep apnea: a systematic review and meta-analysis. Sleep Med [Internet]. 2018;43:100–8. Available from: https://doi.org/10.1016/j.sleep.2017.10.016

34. Archontogeorgis K, Nena E, Papanas N, Zissimopoulos A, Voulgaris A, Xanthoudaki M, et al. Vitamin D Levels in Middle-Aged Patients with Obstructive Sleep Apnoea Syndrome. Curr Vasc Pharmacol [Internet]. 2018 Mar 26;16(3):289–97. Available from: http://www.eurekaselect.com/152736/article

35. Persson LJP, Aanerud M, Hiemstra PS, Hardie JA, Bakke PS, Eagan TML. Chronic Obstructive Pulmonary Disease Is Associated with Low Levels of Vitamin D. Hartl D, editor. PLoS One [Internet]. 2012 Jun 21;7(6):e38934. Available from: https://dx.plos.org/10.1371/journal.pone.0038934

36. Bellia A, Garcovich C, D’Adamo M, Lombardo M, Tesauro M, Donadel G, et al. Serum 25-hydroxyvitamin D levels are inversely associated with systemic inflammation in severe obese subjects. Intern Emerg Med [Internet]. 2013 Feb 25;8(1):33–40. Available from: http://link.springer.com/10.1007/s11739-011-0559-x

37. Urade Y, Hayaishi O. Prostaglandin D2 and sleep/wake regulation. Sleep Med Rev [Internet]. 2011 Dec;15(6):411–8. Available from: https://linkinghub.elsevier.com/retrieve/pii/S108707921100092X

38. Romano F, Muscogiuri G, Di Benedetto E, Zhukouskaya V V., Barrea L, Savastano S, et al. Vitamin D and Sleep Regulation: Is there a Role for Vitamin D? Curr Pharm Des. 2020 Mar 10;26(21):2492–6.

39. Huang W, Shah S, Long Q, Crankshaw AK, Tangpricha V. Improvement of Pain, Sleep, and Quality of Life in Chronic Pain Patients With Vitamin D Supplementation. Clin J Pain. 2013 Apr;29(4):341–7.

40. Majid MS, Ahmad HS, Bizhan H, Hosein HZM, Mohammad A. The effect of vitamin D supplement on the score and quality of sleep in 20–50 year-old people with sleep disorders compared with control group. Nutr Neurosci. 2018 Aug 9;21(7):511–9.

